# Communities facilitating increasing smoke-free homes (CO-FRESH): co-developing an intervention with local stakeholders in Indonesia and Malaysia

**DOI:** 10.1101/2024.10.30.24316337

**Authors:** Rachel O’Donnell, Bagas Suryo Bintoro, Aliya Wardana Rustandi, Retna Siwi Padmawati, Aidil Ikhwan Ahmad, Nur Hanisah Misban, Izzah Athirah Mohd Shu’ah, Nur Shafiqa Safee, Wan Taqiyyah Zamri, Siti Nurhasyimah Ayuni binti Kamni, Aziemah Zulkifli, Isabelle Uny, Emilia Zainal Abidin, Yayi Suryo Prabandari, Sean Semple

## Abstract

**Introduction:** Exposure to second-hand tobacco smoke generates a considerable health burden globally. In south-east Asia, most of that burden falls on women and children who are exposed to second-hand smoke from male smoking in their home. Interventions to encourage smoke-free homes have tended to target smokers individually or within their family unit, although some evidence suggests a community-wide approach holds promise. This study co-developed an intervention toolkit that could be applied to small village/town communities in Indonesia and Malaysia.

**Methods:** The CO-FRESH study used four work packages to: (i) create online training materials to equip local health professionals to tackle smoking in the home; (ii) create a local public information campaign on the benefits of a smoke-free home; (iii) establish methods to provide household air quality feedback to highlight the impact of smoking in the home; and (iv) map local existing services to support families to create smoke-free homes.

**Results:** Four specific communities (two in each country) were involved in intervention development. Training materials for health professionals and a toolkit for use by communities to encourage smoke-free homes were co-created. Communities welcomed the concept of tackling smoking in the home, however there was a lack of knowledge about how second-hand smoke moved around the home and could enter indoor spaces from outdoor smoking. The concept of a ‘smoke-free’ home was often misunderstood, alongside what constitutes indoor versus outdoor space. In addition, findings of high background air pollution levels mean that household air quality measurement may not be suited to providing second-hand smoke information in these communities.

**Conclusions:** Communities in Malaysia and Indonesia recognised the importance of reducing smoking in the home, and welcomed the approach of co-developing community-wide methods of tackling the issue. The CO-FRESH toolkit requires evaluation to determine effectiveness and how it can be implemented at scale.

**What is already known on this topic?:**

- Exposure to second-hand smoke causes nearly 300,000 child deaths per year globally.
- As a result of high prevalences of adult male smoking, children in South-East Asia have a particularly high rates of exposure to SHS; 58% of children are exposed to SHS in Indonesia and 49% in Malaysia.

**What does this study add?:**

- Whilst participants showed some understanding of the health harms associated with second-hand smoke, many did not realise that smoke travels from one room to another, and can linger in the air for up to five hours.
- Although men often accepted smoking in the home as a social norm, and women spoke of challenges raising the issue with them, community members and health professionals welcomed the concept of creating a smoke-free home to better protect familial health.

**How this study might affect research, practice or policy?:**

- This study co-developed the CO-FRESH smoke-free homes intervention toolkit, which could be delivered in small village/town communities in Indonesia and Malaysia by healthcare professionals, community leaders and/or peers, to reduce exposure to second-hand smoke in both countries.

## INTRODUCTION

Almost one-quarter (21%) of Malaysian adults and one-third (31%) of Indonesian adults are smokers.(1,2) In both countries, smoking is predominantly a male behaviour. Forty three percent of men aged 15 years and over are reported to smoke compared with 1.4% of women in Malaysia, with 58.3% of men compared with 3.6% of women smoke in Indonesia.(2,3)

The Malaysian government has adopted a number of measures to reduce the risks of second-hand smoke (SHS) exposure, prohibiting smoking in several types of public places and workplaces,(for example(4,5)) in line with the World Health Organisation Framework Convention on Tobacco Control (WHO FCTC), ratified by the Malaysian government in December 2005. The FCTC encourages signatory countries to implement universal measures to protect non-smokers from SHS exposure, and to ensure at least 90% of their population are protected from SHS exposure through smoke-free policies or laws.(6) However, in Malaysia compliance with smoke-free legislation is reported to be relatively low.(7) This low compliance coupled with the common practice of smoking in the home means that approximately 1 in 4 (25.9%) non-smoking adults report being exposed to SHS in the home, with exposure rates higher among females (31.3%).(8) In a recent study of 420 pregnant Malay women 95% of the 209 who reported being exposed to SHS were exposed at home, as a result of their husband smoking.(9) Few studies have reported on the proportion of children exposed to SHS in the home in Malaysia. A cross-sectional study of 1064 children aged 10-11 years conducted in 2011reported 52.9% of children were exposed to SHS at home.(10) A more recent study used self-report questionnaires with children aged 10-11 (n=312) living in the rural area of Kelantan. The prevalence of children’s SHS exposure at home was 55.8%, with nearly half of children (44%) living in a home with two or more adults who smoked.(1) Only 22% of Malaysian adult smokers and 47% of Malaysian adult non-smokers report having a completely smoke-free home, according to the Global Adult Tobacco Survey (GATS) data.(11)

Indonesia is the only country in the Asia–Pacific region that has not ratified the WHO FCTC. The prevalence of SHS exposure among non-smoking adults in Indonesian homes has been reported as 59.3%(12) The dangers of SHS are not widely recognised and are not commonly taught in medical schools.(13) There is limited data available on the prevalence of SHS exposure in infants and children in Indonesia, however it has been reported that about 57.8% of adolescents aged 13–15 years are exposed to SHS at home.(14)

There is no safe level of exposure to SHS,(15) which has been shown to have wide-ranging adverse health effects on adults and children, including lower respiratory infections and asthma, ischaemic heart disease and lung cancer.(16) Exposure to second-hand tobacco smoke (SHS) causes nearly 300,000 child deaths per year globally, and eliminating or reducing children’s exposure to SHS is a global public health priority.(16–18) Male smoking is particularly common in South-East Asia, and children there have among the highest rates of exposure to SHS (50-60%) in the world. Smoking in the home is also a predictor of child smoking uptake.(19) Smoking at home is declining in the UK and other high-income countries. This is due both to reducing smoking prevalence and changing social norms about smoking in the presence of non-smokers at home. In many LMIC settings, in contrast, there is limited evidence of progress in driving down exposure to SHS in homes.(20) Methods to achieve this have included information provision targeting pregnant women and their partners,(21) delivery through mosques and faith leaders,(22) and providing household-level information of the impact of smoking on air quality.(23,24) These interventions have shown mixed results, which are likely, at least in part, to be associated with broader cultural and social norms, including the social exchange of tobacco as an act of friendship or inclusion in some Asian cultures, and gender-based norms which may cause some women to refrain from challenging men in their household when they smoke in the home.(25)

On this basis, a small number of interventions have adopted a community-level approach to smoke-free home interventions. For example, a programme of research conducted in the rural state of Kerala, India(26) suggests that a 30-60% reduction in smoking in the home may be achieved using a community-wide initiative including education about SHS harms and the establishment of a smoke-free community mandate, backed up by a declaration signed by local leaders. This approach, based on the principle of collective efficacy, seeks to change community smoking norms and highlights the health impacts of SHS exposure in the home on women and children. A similar approach has also been piloted and evaluated in the homes of 296 people who smoke residing in four communities in Yogyakarta, Indonesia.(27,28) Prior to the intervention, 11% of those who smoked did not smoke in their home, this increased to 54% post-intervention. Health educators and community health volunteers were trained to implement smoke-free homes in this intervention, which also focused on smoke-free homes as an issue for women and children, highlighting male responsibility for the welfare of their family. Finally, an intervention providing information on SHS harms, knowledge about support to quit smoking and community-level campaigns delivered in a suburban community in the Pathumthani province in Thailand(29) resulted in a higher proportion of smoke-free homes in the intervention group (75%, n=27), compared with the control group (0%, n=27). The value of community measures to encourage smoke-free homes has also been highlighted in the Kuala Lumpur charter on smoke-free homes, which calls for training of allied-health professionals, greater education in schools about the harms of SHS at home, and improved community support.(30)

This paper reports on the co-development of CO-FRESH (COmmunities Facilitating incREasing Smoke-free Homes) in Indonesia and Malaysia, highlighting lessons learned and adaptations required prior to the development of a future trial to implement and evaluate the intervention.

## METHODS

### a. Development sites

#### Indonesia

Intervention development work was carried out in the Temon Subdistrict, situated within Kulon Progo Regency, west of Yogyakarta. The area is approximately 3,600 hectares in size with a population of 28,000 spread across 15 villages along with 95 sub-villages. The work was centered around the Temon I Public Health Center (PHC), a small inpatient care facility serving eight villages where approximately 50% of the adult population smoke.

#### Malaysia

Intervention development work was carried out in four agricultural settlement areas adjacent to each other under the Federal Land Development Authority (FELDA)(31) in the districts of Kuala Kubu Bharu in the state of Selangor, and Muallim in the state of Perak, approximately 70 kilometers north of Kuala Lumpur. The combined population of these settlement areas is approximately 3,400. These locations were chosen in response to a recent study (32) which reported that 66% of male participants from the rural areas of FELDA in Selangor smoked.

### b. Intervention development

The intervention part of CO-FRESH includes four work packages (WP) (see Table 1). Intervention development in both countries incorporated quantitative and qualitative work with members of the community, including health professionals, community leaders, men who smoke and non-smoking family members.

**Table 1:** CO-FRESH intervention components.

| **Intervention Component** | **Audience** | **Outline/Purpose** |
| --- | --- | --- |
| WP1. Developing online training on smoke-free homes | Health professionals | To increase understanding of the benefits of smoke-free homes, so that health professionals have the tools to provide brief advice at every contact with smokers and their families within the community |
| WP2. Information provision | Public/Local Community | Co-development of local, population level information campaign including a series of posters, word of mouth, social media and media engagement work, to be delivered through workplaces, schools and leisure settings. |
| WP3. Personalised feedback on air quality levels in the home | Families | Low-cost methods to engage local families in citizen science air quality measurement work that will provide personalised data showing how smoking at home impacts on indoor air quality. |
| WP4. Local service mapping and feedback on potential intervention approaches | Families | To identify and document local existing services and potential ways to support smokers and their families to create a smoke-free home. |

#### i. WP1: Online training for local healthcare professionals – module development and sensitisation work

The online training module is designed to be used by health and allied health professionals to deliver a very brief intervention to individuals who smoke and/or their family about the benefits of creating and maintaining a smoke-free home. Research team members in Indonesia, Malaysia and Scotland reviewed existing training materials from Quit Tobacco International’s (QTI) Indonesia smoke-free village module which is designed for use by medical doctors.(33) We adapted these training materials to include peer-reviewed, published findings relevant to both countries and local regions as required, and to make them accessible to a wider range of health professionals. Training on how best to communicate with people who smoke about creating a smoke-free home was also included in this module, based on existing AFRESH intervention materials, which were previously developed by members of the team.(34) Following initial adaptation, discussions were held with relevant health professionals in both countries to ascertain their perceived confidence in delivering the module, suggested improvements, additional resources or support that would be required to enhance the effectiveness of module delivery, and their views on the relevance/importance of the module. Within this WP, sensitisation work was also undertaken to involve the local community through engagement meetings with community leaders and local stakeholders, to build support and understanding for this work from the outset. Findings from this sensitisation work fed into development of the WP2 toolkit to maximise it’s acceptability.

#### ii. WP2: Development of information campaigns for the local population about SHS harms

A series of focus group discussions were held in Malaysia and in Indonesia, with men who smoke, non-smoking adult household members (male and female), and community leaders (male and female). One purpose of these focus groups was to shape the development of a series of locally acceptable key messages to communicate the harms associated with SHS and the benefits of creating a smoke-free home. To inform the development of these key messages, participants were shown a range of statements delveoped by the team (See Supplementary file 1), and asked to identify whether each statement was a myth or a truth. General views on/reactions to each statement were also noted. Information was also gathered to shape the development of a toolkit of local materials including posters, videos and social media content, which could be delivered across multiple platforms in the future, and through workplaces, schools and leisure settings. To facilitate the development of culturally-appropriate, acceptable materials, views were elicited on preferred terminology (e.g. SHS exposure or passive smoking).

#### iii. WP3: Personalised feedback on air quality levels in the home

Drawing on previous projects (the AFRESH intervention(34) in the UK and the MyFamilyMySmoke project(8,23) in Malaysia), we tested the feasibility of engaging families in a low-cost method of indoor air quality measurement, which enabled provision of personalised data showing how smoking in the home home produces high indoor air pollution concentrations and how these compare to pollution from other sources. In each country, families living in the local community were provided with a participant information sheet, and after 48 h, with consent to participate provided, a suitable date for in-home air quality measurement was agreed.

Using previously published methods,(23) 7-day measurement of fine particulate matter (PM_2.5_) concentrations in the home were made using PurpleAir PA-II-SD air quality monitors. Monitors were delivered to the home, with written instructions on self-installation in the main living area, at least 1m from the ground and from any doors/windows. Mean PM_2.5_ concentrations were calculated, and readings were used as a means of providing personalised feedback on air quality levels in the home. This feedback was provided at the start of a group interview conducted with household members after the 7-day measurement period. Each interview explored views and reactions regarding the personalised air quality feedback received, the extent to which receiving personalised indoor air quality feedback might impact on home smoking behaviours, and whether any improvements could be made to the delivery and/or content of feedback received (see Supplementary file 2 – interview schedule).

#### iv. WP4: Local service mapping and feedback on potential intervention approaches

As part of any future community-wide smoke-free homes intervention delivery programme, it will be essential to provide individual support to those who wish to change their own smoking behaviour – either by making their home smoke-free by smoking outside, or by quitting smoking. On this basis, focus group discussions included a question on whether people who smoke were aware of the existence of local smoking cessation or smoke-free home support services, which the team documented as a mapping exercise. Participants were also asked for their views on a range of potential community-wide smoke-free home interventions, including the use of a household declaration, as employed in previous work conducted by team members in Indonesia,(35) and Malaysian’s Smoke Free Home (Rumah Bebas Asap Rokok or RBAR) and MyHOUSE approaches.(36) This ensured that any cultural and/or societal differences identified were taken into account in both countries during intervention development.

### c. Analysis

All focus group discussions and interviews were transcribed, anonymised and uploaded into NVivo 12 for coding. Transcripts were coded by two members of each team, and then analysed using the framework approach(37) alongside use of memos to support reflexivity. (38) A thematic framework was developed by the team to guide data analysis, using deductive (considering the topic guide) and inductive (reading transcripts and coding) techniques. Data summaries were written in relevant cells of the framework grid, and these were used to identify high level themes before further in-depth analysis was conducted. Themes were finalised based on re-examining data and reflexive team discussions. For the air quality data, two-minute concentration values in micrograms/m^3^ (μg/m^3^) of PM_2.5_ were extracted from the onboard SD card on the PurpleAir device and analysed in MS Excel. For the entire duration of measurement in each home the mean and median concentrations were calculated. The 2-minute maximum and minimum values recorded over the sampling period were also extracted.

### d. Community engagement and involvement

In both countries, we ensured that relevant health professionals, community leaders and/or members of the public were adequately involved in each stage of intervention development. As outlined above, health professionals helped to shape the content of the draft training module and community leaders inputted to the development of the toolkit to ensure acceptablity of key messages (WP1). Members of the public shaped the finalisation of key messages contained within the toolkit (WP2). They also provided views on the personalised air quality feedback received (WP3) to shape feedback delivery style and content, and on the cultural acceptability of various community-wide smoke-free home interventions prior to future intervention development (WP4).

### e. Ethics

The study was granted ethical approval by University of Stirling General University Ethical Panel (GUEP 2022 10527 7993, 25/10/2022), Universitas Gadjah Mada Faculty of Medicine, Public Health and Nursing Medical and Health Research Ethics Committee (KE/FK/0099/EC/2023) and the University Putra Malaysia Ethics Committee for Research involving Human Subjects (JKEUPM-2023-047).

## RESULTS

### a. WP1: Module development - outcomes

The online training module on SHS and the benefits of creating a smoke-free home was generated for health and allied health professionals. The training was organised into four sub-modules comprising presentations, handouts and/or videos. Each section concentrated on particular knowledge bases and the steps to assist community members in implementing a smoke-free home. Each sub-module was supplemented with a video and more detailed information to enhance knowledge knowledge (see Supplementary file 3).

To ensure participants achieve learning objectives, tests are incorporated at the beginning and end of each section. The training module is available via an online learning platform (e-LOK) hosted at the University of Gadjah Mada.

The modules underwent pre-testing through audio-recorded interviews (see Supplementary files 4 and 5 for interview questions) with a total of eleven health professionals (six from Indonesia and five from Malaysia) drawn from the communities where we were piloting the programme. Respondents noted that the content was comprehensive and delivered concepts in language that was easily understandable: the accessibility making it suitable for both medical and non-medical personnel, and respondents reported no issues accessing it through e-LOK. Some feedback from participants focused on editorial improvements, suggesting the inclusion of illustrations to enhance understanding. Additionally, there was a demand for both online and offline (handout) versions of the health education media. Respondents stressed the importance of emphasizing frequently asked questions within the module, as it is expected to provide theoretical and technical answers to common queries encountered in the field. These suggestions were incorporated to further enhance the usability and effectiveness of the module for its intended audience.

### b. WP2 findings

The composition of each community focus group is summarised in Table 2 for each country. Key findings are summarised in Table 3.

**Table 2:** Focus group composition by country.

| **Focus group** | **Composition** | **Gender** | **Smoking status** |
| --- | --- | --- | --- |
| Indonesia |  |  |  |
| 1 (N=10) | Community members | Female | Non-smokers |
| 2 (N=11) | Community members | Male | Smokers |
| 3 (N=8) | Community stakeholders | Male | 3 ex-smokers,<br>1 smoker<br>4 non-smokers |
| Malaysia |  |  |  |
| 1 (N=3) | Community leaders/worker | Female | Non-smokers |
| 2 (N=4) | Community leader and members | Male | Smokers |
| 3 (N=3) | Community leaders | Male | Non-smokers |
| 4 (N=2) | Community members | Male | Smokers |
| 5 (N=3) | Community leaders | Female | Non-smokers |
| 6 (N=3) | Community leader and members | Female | Non-smokers |
| 7 (N=7) | Community leader and members | Female | Non-smokers |
| 8 (N=5) | Community members | Male | Smokers |

**Table 3:** Key focus group findings.

| Topic guide area | Key findings | Example quotes |
| --- | --- | --- |
| Health harms associated with SHS exposure | Participants generally recognize the harmful effects of smoking indoors. | *"We already know the dangers, right? The dangers of smoking, it causes various diseases..." (Indonesia, FG3)*<br>*"A [cigarette] stick or half a stick it is still a danger to others...babies and all" (Malaysia, FG3)* |
| Preventing SHS exposure in the home | Some participants believe that quitting smoking is most effective way to create a smoke-free home | *"I agree that the only way [to create a smoke-free home] is to stop smoking. Because even when we smoke in open spaces, the smoke will get on our clothes and enter the house." (Indonesia, FG3)*<br>*"If you don't want to have smoke exposure in the house, you have to stop smoking."*<br>*"I think so too. No smoking in the house. So those who smoke will have to stop." (Malaysia, FG4)* |
| How SHS travels | Many participants are unaware that 85% of SHS is invisible, and that it can linger in the air for up to 5 hours | *"...Cigarette smoke can move from one room to another. For example, if someone smokes on the terrace, it can reach the living room, reach the bedroom, right? That means it's correct. But as for the 85%, I don't know, I've never calculated it, but...I thought it disappeared, but it turns out it sticks invisibly, and I think the percentage depends on the condition of the house. If the house is closed, with air conditioning, then maybe it can be 100%, if it's open, it's less." (Indonesia, FG1)*<br>*"For me...it's not that the smoke stays...smoke is very fine? For example, even if you smoke in the bathroom, the bathroom may not be completely closed. There must be some holes that smoke can escape." (Malaysia FG4)* |
| Indoor vs outdoor air pollution | Participants often perceived outdoor air pollution as more harmful to health than indoor sources, including when smoking takes place in the home. | *"I would like to add that cigarette smoke is specifically intended for... especially when we are indoors, the smoke is for ourselves. But as for air pollution outdoors, it comes from various sources like dirt, vehicles, and so on. It all mixes together, making the air more polluted. Inhaling this polluted air is actually more dangerous and can lead to diseases." (Indonesia, FG2)*<br>*"If it's industrial [outdoor air pollution], the smoke, chemical composition...we don't know if it's more than cigarette smoke". (Malaysia, FG3)* |
| General beliefs about smoking | Men often accepted smoking as a social norm while women prioritized their health by avoiding exposure to SHS. | *"...But with these friends, I just try to socialize with them, it's normal solidarity. They smoke but I don't, and that's ok."* (Indonesia, Male, FG3)<br>*"So I have to be careful and avoid situations where there are gatherings with people smoking. It's better for me to protect myself, use a mask or stay away, just avoid cigarette smoke"* (Indonesia, Female, F1)<br>*"From there ... from their children to grandchildren, they all watched them, they followed, and smoked cigarettes too"* (Malaysia, Male, FG2)<br>*"Even I don't have asthma, but I can't stand it [being exposed to SHS]"* (Malaysia, Female, FG7) |
| Talking to other family members about SHS exposure | Women spoke of challenges raising the issue of smoking in the home with men. | *"The difficult part is that I'm afraid to tell them [I don't like being exposed to SHS] because I don't want to offend or upset them."* (Indonesia, FG1)<br>*"Not taking heed [of where smoking takes place] is why sometimes uh that's the cause of the quarrel"* (Malaysia, FG5) |
| Avenues for future behaviour change interventions | Men expressed a willingness to smoke outside rather than in the home. | *"If we realize, 'Oh yes, smoking inside the house can make the wife angry', then we are aware of it and smoke outside."* (Indonesia, FG2)<br>*"Sometimes even us smokers, do not want to smoke in the house, we will go out automatically unless living alone"* (Malaysia, FG4) |
|  | Participants emphasized the importance of providing visible reminders not to smoke indoors, along with designated outdoor smoking areas to accommodate smokers. | *"Well, if there's a hangout place down the street, that [smoking outside] might be possible. But if there's no place to hang out, it's difficult, it's not polite to ask people to smoke while standing [outside]. Because they are guests."* (Indonesia, FG1)<br>*"[In designated outdoor smoking areas] we can also try to put up posters...for example on the road with the signboard...what are the dangers of smoking, do not smoke cigarettes [indoors].."* (Malaysia, FG7) |
|  | The absence of ashtrays and lighters indoors were suggested as effective deterrents to smoking in the home. | *"Usually, smokers carry lighters, but to prevent it [smoking in the home], we shouldn't provide lighters or ashtrays in the house."* (Indonesia, FG1)<br>*"Me...my advice is easy if it's my house, I will not set a place to smoke, so when people come, they will see on the table and there is no ashtray, so I think they will not smoke. So, they will think, uh, the landlord doesn't smoke. This is my opinion so to encourage people not to smoke in my house, I will not set a smoking place."* (Malaysia, FG7) |
|  | Some participants suggested it would be unusual to smoke outdoors alone, or during inclement weather. | *"If we want to smoke outside, let's say we go out to the terrace but [smoking] alone, it's like a lost person [laughs]."* (Indonesia, FG2)<br>*"It depends on the weather, whether it rains or not. If it doesn't rain, we can do it [smoke outside the house]"* (Indonesia, FG2) |
|  | Family agreements were seen as crucial in establishing and maintaining a smoke-free home. | *"If it's your own family, there must definitely be an agreement, meaning we may create some rules. For example, if someone violates the rules, there should be a punishment, and if they follow the rules, there should be a reward."* (Indonesia, FG3)<br>*"whoever comes to my house doesn't matter guests or my siblings ... who smokes, he does come to my house whether it's Eid or what ... smoke cigarette outside. I will not allow smoking in the house".* (Malaysia FG7) |
|  | Women suggested that guests/visitors should be reminded not to smoke in their home, but that this can be challenging. | *"The intention is to forbid smoking, but in a subtle way. 'You can smoke, but outside.' If we often speak like that, 'Oh, this house doesn't want cigarette smoke, there are small children here,' they will feel uncomfortable smoking."* (Indonesia, FG1)<br>*"I think it's a little difficult... My view is like that if it's family maybe we can control [ask them not to smoke in the home], but in this village, for me it's quite difficult to prevent people from smoking."* (Malaysia, FG7) |
|  | Participants suggested that tighter government regulations would help support community smoke-free initiatives. | *"If smoking is to be tightened, then the government plays a big role. There must be legal regulations."* (Indonesia, FG1)<br>*"Only the enforcement can do this." "It's not wrong that the government made some effort but it is going to take time to get the results"* (Malaysia, FG4) |

In relation to the truths and myths presented to each group, respondents generally recognized the harmful effects of smoking indoors, even a single cigarette. However, some participants believed that quitting smoking is the only effective means of creating a smoke-free home, neglecting the alternative of smoking outdoors, and most misunderstood how SHS travels within indoor spaces. Many participants were unaware that around 85% of cigarette smoke is invisible, meaning they are likely to underestimate level of exposure and the duration of exposure when SHS is present. Thirdly, respondents often perceived outdoor air pollution concentrations as being higher and more harmful than indoor pollution even when smoking takes place in the home.

Gender disparities in terms of smoking and beliefs were apparent in both countries, with men often accepting smoking as a social norm while women prioritized their health by avoiding exposure to SHS. Additionally, there was reluctance among non-smokers to confront smokers, particularly in Javanese culture, due to fear of causing offense, especially when the smoker holds a higher status.

Interestingly, (male) smokers expressed a willingness to be reminded to choose their smoking locations wisely, suggesting potential avenues for behaviour change interventions. Measures only permitting smoking on terraces were met with scepticism, as individuals suggested it would be peculiar to smoke alone outside, especially during inclement weather. The participants brought up several strategies for promoting smoke-free environments within households and communities. Participants emphasized the importance of providing visible reminders not to smoke indoors, along with designated outdoor smoking areas to accommodate smokers. Additionally, the absence of smoking facilities such as ashtrays and lighters were suggested as effective deterrents. Family agreements regarding smoking were seen as crucial in establishing and maintaining a smoke-free home. Discussions also centred on how hosts had to remind guests not to smoke indoors, particularly if vulnerable individuals or children were present or if the host themselves did not smoke. Participants also raised the concept that government regulations would help support smoke-free initiatives on a larger scale within communities.

### c. WP3: Personalisation – outline PM_2.5_ levels, comment on perceived acceptability of air quality monitoring and reactions to results

Purple Air II devices were installed in a total of 12 houses in Indonesia and 10 in Malaysia to capture data on household PM_2.5_ concentrations. All of the homes included an adult who smoked at home. Other potential sources of PM_2.5_ included cooking, mosquito coil burning, vehicle (motorcycle/moped) engines, and burning tyres for bicycle tyre repair was noticed.

Table 4 shows the duration of measurement, median, minimum and maximum values measured in each home in both Indonesia and Malaysia.

**Table 4:** Duration of measurement, median, minimum and maximum PM_2.5_ values measured in each home.

| Home | Duration of sampling (days) | Mean PM<sub>2.5</sub> (µg/m<sup>3</sup>) | Median PM<sub>2.5</sub> (µg/m<sup>3</sup>) | Minimum PM<sub>2.5</sub> (µg/m<sup>3</sup>) | Maximum PM<sub>2.5</sub> (µg/m<sup>3</sup>) |
| --- | --- | --- | --- | --- | --- |
| **Indonesia** |  |  |  |  |  |
| 1 | 8 | 57 | 32 | 3.6 | 1036 |
| 2 | 8 | 89 | 33 | 3.0 | 1737 |
| 3 | 7 | 55 | 33 | 3.4 | 943 |
| 4 | 7 | 62 | 35 | 4.4 | 1351 |
| 5 | 7 | 42 | 35 | 2.4 | 290 |
| 6 | 7 | 43 | 29 | 1.9 | 783 |
| 7 | 7 | 37 | 28 | 2.2 | 508 |
| 8 | 7 | 64 | 44 | 7.3 | 576 |
| 9 | 7 | 53 | 38 | 7.0 | 936 |
| 10 | 7 | 46 | 31 | 6.6 | 712 |
| 11 | 8 | 51 | 35 | 1.7 | 836 |
| 12 | 8 | 72 | 43 | 0.8 | 1212 |
| **Malaysia** |  |  |  |  |  |
| 1 | 7 | 32 | 22 | 8.8 | 1098 |
| 2 | 6 | 39 | 27 | 10.5 | 1071 |
| 3 | 7 | 29 | 23 | 9.0 | 314 |
| 4 | 7 | 18 | 14 | 5.8 | 305 |
| 5 | 7 | 32 | 21 | 8.2 | 252 |
| 6 | 6 | 16 | 14 | 6.0 | 316 |
| 7 | 7 | 34 | 26 | 7.9 | 469 |
| 8 | 7 | 43 | 31 | 20.5 | 344 |
| 9 | 7 | 36 | 30 | 16.4 | 560 |
| 10 | 7 | 28 | 23 | 8.9 | 168 |

The findings on personalized air quality feedback (see Figures 1a and 1b for an example of the feedback provided to each participant) highlighted a range of responses from participants. Women predominantly reacted with surprise and concern upon receiving results, reflecting worries about potential health impacts associated with smoke exposure within their homes. Conversely, men exhibited a mix of emotions, including feelings of guilt, surprise, and in some cases, a sense of anticipation regarding the likelihood of high readings. Many participants speculated that the elevated PM_2.5_ measurements were possibly attributable to activities such as garbage burning in the vicinity rather than smoking behaviour in the home.

**Figure 1a:**
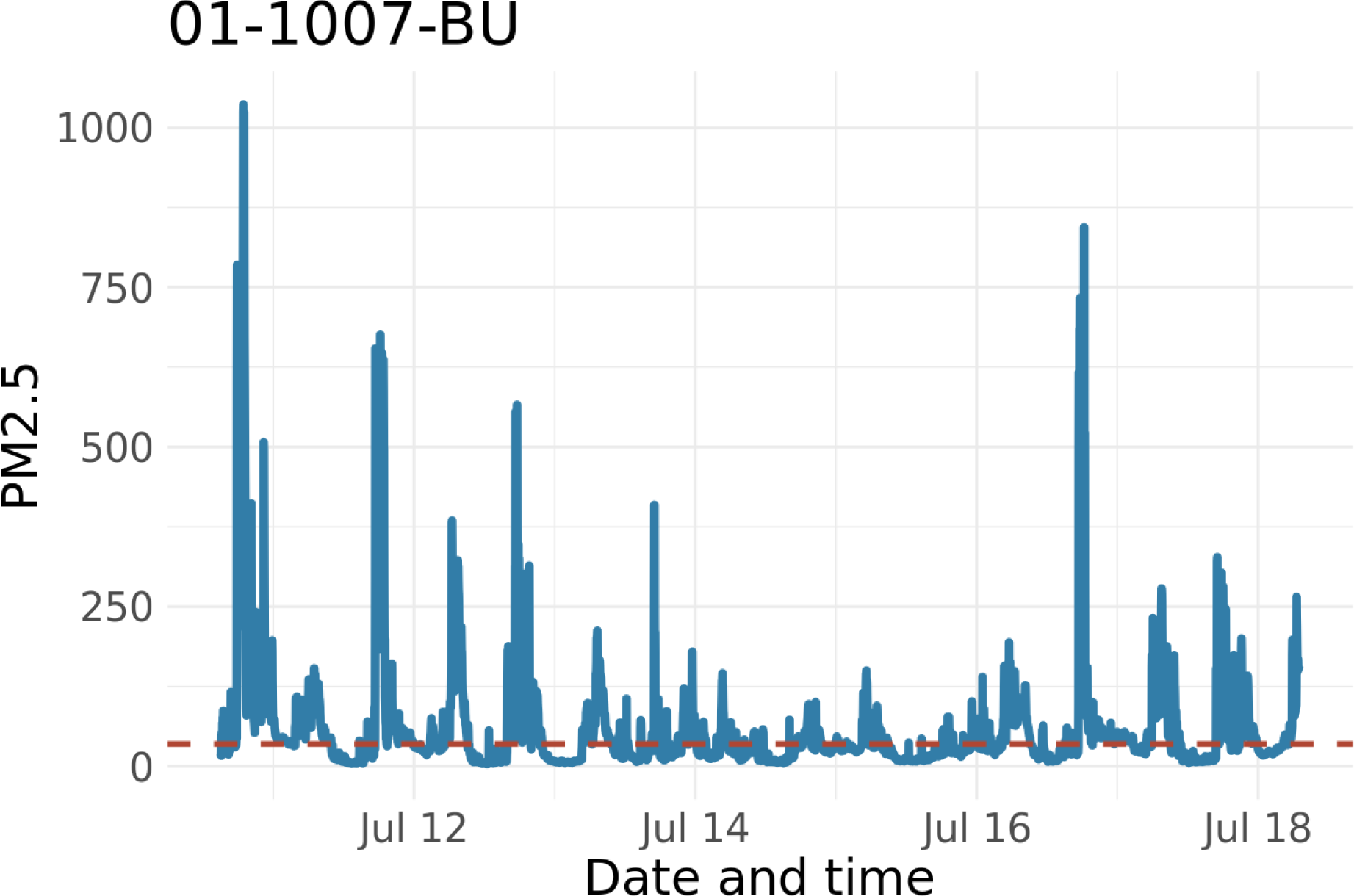
Example measurement of PM_2.5_ concentrations from one home over the ourse of 7-days.

**Figure 1b:**
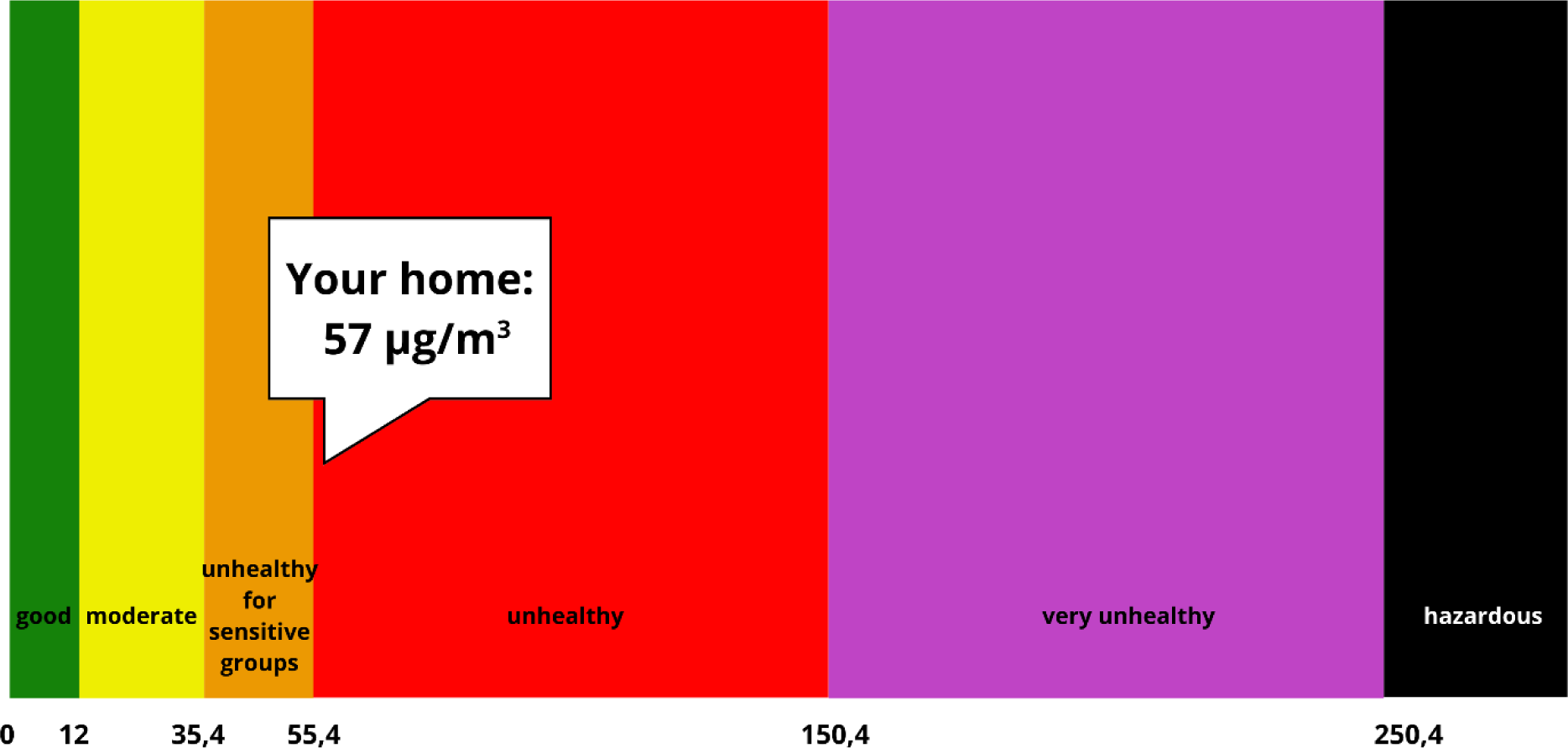
Color-coded graph using the PM_2.5_ standard from WHO for the average PM_2.5_ level during the measurement period.

In response to the feedback received, participants commonly engaged in discussions about the results with their spouses and occasionally extended these conversations to include close neighbours or family members. Some individuals expressed a sense of satisfaction with the insight provided by the measurement device on the impact of smoking on the air quality within their homes.

Regarding potential changes in smoking behaviour prompted by the feedback, there were indications that some participants may make efforts to adjust their habits, such as opting to smoke outdoors after becoming aware of the measurement results. However, transitioning to a completely smoke-free home remained a significant challenge for many participants. Participants widely acknowledged the harmful effects of cigarette smoke, yet not smoking in the home remained a formidable challenge for smokers.

Feedback regarding the provided information revealed that while some participants struggled to fully comprehend the data without additional explanation, others demonstrated a basic understanding, particularly regarding the significance of colour changes (PM_2.5_ concentration changes from one US EPA health band to another) in the feedback.

### d. WP4: Availability, awareness and uptake of of smoking support services

As part of the focus group discussions in WP2 in each country, awareness of local smoking cessation services was explored. In both countries there was a lack of awareness of the existence of these services and a notable reluctance among smokers to engage with these services. Smokers often reported waiting until faced with health issues before considering cessation.

WP4 also involved some simple mapping of services to help smokers and families either quit smoking or create a smoke-free home. In Indonesia, Primary Health Centres are the local health hubs for delivery of smoking cessation services. Primary Health Centres suffer from limited staffing, hindering their capacity to provide comprehensive smoking cessation support. Despite efforts to train health professionals as counsellors, the uptake of smoking cessation services remains minimal in Indonesia and there is no dedicated focus on assisting smokers to create a smoke-free home. In terms of resources for smoking cessation services, significant gaps were also identified. For instance, there is a notable absence of tools to measure objective biomarkers of smoking activity such as exhaled carbon monoxide (eCO) levels, a crucial component in monitoring and managing smoking cessation efforts.

In Malaysia there are several smoking cessation and smoke-free support services available. Examples of support services include smoking cessation clinics, quitline services and community-based programs. Many public and private hospitals and health clinics offer smoking cessation clinics which provide counselling, behavioural support, and medication such as nicotine replacement therapy or prescription medications to help individuals quit smoking. Moreover, the Malaysian Ministry of Health operates the Quitline Malaysia service (jomquit.com), which provides free telephone counselling, advice and support for individuals who want to quit smoking. Non-governmental organizations (NGOs), community centres, and local health departments may organize community-based programs such as educational workshops, awareness campaigns and events. They engage with communities, workplaces, and schools to promote smoke-free environments, raise awareness about the harms of smoking and SHS, and encourage smokers to seek help for quitting. KOSPEN which stands for Komuniti Sihat, Pembina Negara (Healthy Community, Nation Builder), is a community-based health intervention program in Malaysia, aiming to engage with communities to raise awareness about the harmful effects of smoking and SHS exposure. The program also encourages households to adopt smoke-free policies. This includes promoting the benefits of having smoke-free homes, such as protecting family members, especially children, from the dangers of SHS.

Despite the availability of these services in Malaysia, our focus group participants reported that there is minimal uptake or engagement with these services. Smokers may recognize the importance of quitting but face various barriers that prevent them from fully committing to cessation programs or attending support sessions regularly. Some individuals were not aware of the availability of smoking cessation/smoke-free home services or did not know how to access them.

## DISCUSSION

This study co-developed a smoke-free homes intervention toolkit that could be applied to small village/town communities in Indonesia and Malaysia. Interview findings with healthcare professionals suggest that the toolkit training module is easy to understand, accessible and was useable. Focus group discussions with community leaders and household members highlight significiant misunderstandings regarding: how SHS travels within indoor spaces; effective methods of creating a smoke-free home; and misperceptions regarding the comparative risks of exposure to SHS in the home compared with exposure to outdoor air pollution. Whilst women priotirise their health by avoiding exposure to SHS in the home where possible, men often accept smoking in the home as a social norm. Focus group discussions also highlighted a reluctance to raise the issue of smoking in the home with smokers for fear of confontation. Collectively, these findings highlight the need for increased education and support to reduce exposure to SHS in the home in both countries, and the developed toolkit has an important potential role to play in this regard. The CO-FRESH toolkit provides an integrated approach to tackle the issues of SHS in homes in small communities. The co-development of this intervention ensures that it does so in a manner that is ‘bottom up’ through involving smokers, their families, local health professionals and community leaders.This builds on previous work in Indonesia which was ‘top down’ in providing villages with pre-created intervention materials.(35)

### Why use the village unit to encourage smoke-free homes?

Smoke-free home interventions focused on individual or household-level behaviour change may have limited value in South-East Asian settings. In previous research conducted in Indonesia, women expressed a low sense of self efficacy in individually getting their husbands to quit smoking in their homes, but a strong sense of collective efficacy that husbands might agree to a well-publicized community smoke-free homes initiaitve. Men and women expressed concern about the social risks associated with asking guests not to smoke in their homes without a community-wide initiative and visible displays communicating their participation in this movement.(42) In our study, we found that men tend to accept smoking as an important part of socialising, indicating that smoking is still an acceptable norm. It has been suggested that establishing smoke-free homes as a new social norm requires consensus-building endorsed by all residents in a community, redefining SHS exposure as a health issue of women and children and tying family welfare to core cultural values linked to male responsibility to protect the health of women and children.(25,35)

### The need for a common understanding of terms and language relating to smoke-free homes

Understanding what constitutes a smoke-free home in terms of the areas where smoking should not take place and where the boundary lies between outdoors and indoors was variable across our study participants, and reflects previous work in Scotland. In addition, some participants considered that ‘smoke-free home’ meant a home where no-one who smoked lived (i.e. all people in the home were non-smokers). The extension of this was that people believed that smoking cessation was the only method possible to provide their families with a smoke-free home. This is similar to some of the findings from previous qualitative research in Scotland shortly after implementation of smoke-free public spaces legislation.(39)The findings suggest that the definition of smoke-free homes varies between individuals and households. There is a clear need for education and public information that a ‘smoke-free home’ does not have to mean a ‘smoker-free household’.The CO-FRESH toolkit makes this clear for future users.

### Emphasis on childhealth: pros and cons

In this research, air quality feedback and how smoking potentially modified fine particulate matter (PM_2.5_) concentrations in the air, shocked the parents and often focused their attention on their children’s health. This builds on findings from several other countries where feedback of SHS concentrations shocked and motivated parents to consider the impact of their indoor smoking.(40,41) In Bangladesh, many individuals believed that the significance of having a smoke-free home is to improve their child’s health, and this is often further enhanced by children’s direct requests to adults not to smoke.(42)

Although emphasising child health may be a useful method to assist fathers in changing their home-smoking behaviour, the intervention materials should make sure that they underline the definition of a smoke-free home and the need to protect all children in the home. Previous work has suggested that over 60% of smoking fathers reported that they still smoked at home when their children were not present as they believed that the smoke would disappear before their child returned and that this would still classify as providing a smoke-free home.(43)This lack of knowledge about how long SHS remains in the air is a common misconception and is a key part of the education provided in the CO-FRESH toolkit. Previous studies conducted in high-income countries have suggested that when children in the household mature, the strictness of enforcing smoke-free rules at home often eases. The return to smoking in proximity to children occurs gradually and without formal discussion or negotiation.(44,45) This tendency to relax restrictions as children become older, even though all children should be protected from SHS in the home, is also addressed in the CO-FRESH toolkit.

### Strengths and weaknesses

The development of CO-FRESH was carried out in small rural villages or communities and may not be useful or generalisable to more urban settings where community cohesion is less well established. In Indonesia the co-development of the intervention was conducted in suburban Kulonprogo, Java, an area currently experiencing dynamic changes due to development, such as an international airport nearby. This context-specific dynamic may limit the generalizability of the findings to other regions across Indonesia or south-east Asia. This research also focused on the development of the smoke-free home intervention and did not evaluate the roll-out or effectiveness of the intervention itself.

A recent review highlights that the majority of studies on smoke-free homes have taken place in high-income countries.(44) One key strength of our study is that intervention development took place in Malaysia and Indonesia, representing middle-income and low-income countries. Our focus on community engagement and involvement in the co-development of CO-FRESH is likely to have a positive impact on the effectiveness of the intervention,(46) which now requires testing.

## Conclusion

This study successully co-developed a smoke-free homes intervention toolkit that could be applied to small village/town communities in Indonesia and Malaysia.There is now a need to pilot test CO-FRESH in a range of different communities in both countries, to assess whether the intervention effectively reduces children’s exposure to SHS, and determine the contextual factors which maximise recruitment and effective intervention delivery.

## Supporting information

Reflexivity statement

## Data Availability

All data produced in the present study are available upon reasonable request to the authors.

