## Supplementary file 1 for "Communities facilitating increasing smoke-free homes (CO-FRESH): co-developing an intervention with local stakeholders in Indonesia and Malaysia"

**Supplementary file 1: CO-FRESH WP2 Topic Guide**

**Welcome and Introduction (5 mins)**

- Thanks for joining us this today. I’m going to start by giving a quick overview of the research, why we are here, and we’ll have some quick introductions before we start
- This study is about smoking in the home. We’re keen to hear your thoughts on a range of messages and ideas that we share with you. You might not agree with everything you hear and your views may differ – that’s ok. If you have a thought or idea, please do share it.

You’ve all received the information sheet and the consent form. But, I’ll just give a quick overview of the key points around participating:

- The session will last no longer than 1 hour 30 minutes
- It’s a confidential discussion (but we are in a group, so don’t share anything you wouldn’t want others to know) Please only share what you’re comfortable with.
- If you need a break, please take one
- Respect each others views – we only have one person speaking at a time
- Does anyone have any questions?
- If anyone has any issues/questions as we go, please let me know.

**Introductions:** Before we begin, we’d like everyone in turn to very briefly tell us your name and:

(FOR THOSE WHO SMOKE/NON-SMOKING HOUSEHOLD MEMBERS) your role in the household (i.e. parent, grandparent, aunt, uncle)

(FOR COMMUNITY LEADERS) your role in the community

For example my name is XX and my role in this household is XX (ask everyone in turn to introduce themselves).

**Section A: Understanding of SHS and smoke-free home (15 mins) (ALL SECTION A QUESTIONS ARE FOR EVERYONE)**

1. **Firstly, which of these terms do you recognise – ‘second-hand smoke’ or ‘passive smoke’?**

- Show a photo/image of what we mean by second-hand smoke to assist with this discussion
- Establish whether the group is familiar with one or both terms and continue to use the term they are most familiar with throughout this discussion.

2. **What do you understand by the term second-hand smoke/passive smoking?**
   a. *Ask people to share their views with the group*
   b. *Identify any similarities/differences between responses*
   c. *Where do these understandings come from? What influences them?*
3. **What do you understand about any harms related to second-hand smoke/passive smoking?**
   a. *Ask people to share their views with the group*
   b. *Identify any similarities/differences between the group. Probe for understanding regarding health harms, and harms for specific groups (children, non-smokers, pregnant women, older adults, smokers)*
   c. Where do these understandings come from? What influences them?
   d. Do you have any concerns or worries about second-hand smoke? For example, are you more concerned about health risks or the smell, and why? Do you have any other concerns about second-hand smoke?
4. **What do you understand by the term ‘smoke-free home’?**
   a. *Ask everyone to share their views with the group*
   b. *Identify any similarities/differences between the group, for example if smoke-free home means ‘smoker-free home’ to some people* (explain that this is not what we mean and why)
   c. How achievable is it for you to create a completely smoke-free home, where smoking only takes place outdoors? Explain that a completely smoke-free home includes not smoking on terraces or balconies as smoke can drift back indoors. Explore responses briefly, including any challenges raised – do others feel the same, or differently?

**Section B: Myths and Truths about second-hand smoke (25 minutes) (ALL SECTION B QUESTIONS ARE FOR EVERYONE)**

[Each statement printed on laminated card - with the ‘myth’ on Side A and the ‘truth’ on Side B]

We’re going to show you a series of statements about second-hand smoke and smoking in the home, and we’d like to hear your views on them. Some are true, and some are false – can you tell us which ones you think are true and false? (don’t give titles here – they are for our reference only)

**For each myth and truth card:**

- Wait until everyone has expressed their view, and then explain which the myth is and which the truth is.
  - Do you agree that the ‘truth’ statement is fact? Why?
  - You seem [shocked, unsurprised] that this is true. Why is that?
  - Do you agree that the ‘myth’ statement isn’t true? Why?
  - You seem [shocked, unsurprised] that this isn’t true. Why is that?
- Answer any related questions that participants might have and if applicable, address any concerns expressed, before moving onto the next pair of messages.

a. **Preventing SHS exposure in the home**

MESSAGE 1 (MYTH): The only way to prevent second-hand smoke exposure in the home is to stop smoking completely.

MESSAGE 2 (TRUTH): You can minimise second-hand smoke exposure by not smoking in the home, and smoking outdoors only.

b. **How second-hand smoke travels**

MESSAGE 1 (TRUTH): Second-hand smoke can move around the home from room to room and 85% of second-hand smoke particles are invisible.

MESSAGE 2 (MYTH): Smoking in one room such as the bathroom, with the window open/door closed, means that second-hand smoke will not affect family members elsewhere in the home

c. **Indoor versus outdoor pollution**

MESSAGE 1 (MYTH): Outdoor air pollution is more of a problem than second-hand smoke in the home

MESSAGE 2 (TRUTH): Second-hand smoke can be just as damaging to health as outdoor air pollution, if not worse.

d. **How many cigarettes can you smoke in the home before it becomes harmful?**

MESSAGE 1 (TRUTH): Smoking even just one cigarette in the home can cause harm to others: there is no safe level of second-hand smoke in the air.

MESSAGE 2 (MYTH): Just having one or two cigarettes in the home is not harmful

e. **Smoking on your own in the home**

MESSAGE 1 (MYTH): Smoking in the home when no-one else is around stops family members from breathing in second-hand smoke.

MESSAGE 2 (TRUTH): Second-hand smoke lingers in the air for several hours, so waiting until no-one else is in the home may not fully protect others from second-hand smoke exposure.

**Section 3: Interventions (40 minutes)**

**For those who smoke:**

1. Has anyone ever spoken to you about not smoking in the home? If so, who, and was this helpful? If not, would this be helpful? Who do you think should speak with you about this (prompt: health clinic staff, family members, community members etc)?

**For non-smoking family members:**

2. Have you ever spoken with family members who smoke about them not smoking in the home? If so, who, and did this help or not? Why? Who do you think should speak with XX [family member] about this (prompt: health clinic staff, family members, community members etc)?

**For community members/leaders:**

3. Has you ever spoken with anyone about smoking in the home? If so, who, and did this help or not? Why? Who do you think should speak with people who smoke about this (prompt: health clinic staff, family members, community members/leaders etc)?

**QUESTIONS FOR EVERYONE: (ask each question ‘for you’, ‘for your family’ or ‘for your community’ depending on participant type)**

We don’t really know the most effective ways to help families to create and maintain a smoke-free home. We’d like to hear your views on a few possible options, because this can sometimes be difficult to achieve:

4. In some parts of the UK, people are advised to ‘take 7 footsteps out of the home to reduce second-hand smoke exposure. What do you think of this idea? Could this work a) for you b) for your family c) for others in your community? Why/why not?
5. Some studies have suggested that providing households with personalised information about the impact of smoking on air quality in the home could help households to make changes to reduce smoking in the home. This involves use of an air quality monitor, a small machine which you plug in and place in your front room for a few days at a time (have an air quality monitor with you to show people, and an example graph as well to show the feedback that household members receive). Is this something that you think would interest a) you b) your family c) this community or not? Why? (Note that it costs next to nothing to run)

**QUESTION FOR THOSE WHO SMOKE ONLY (MALAYSIA):**

6. Have you ever used nicotine replacement therapy before? (show picture examples, patches, gum, inhalator etc) Is this available from your local health clinic? Some studies have suggested that using nicotine replacement products (i.e. gum, lozenges, mouthspray) when you are indoors (instead of smoking indoors) could help people to create/maintain a smoke-free home and reduce smoking. Nicotine replacement products tend to be used to support people to quit smoking completely, so what are your views on use of nicotine replacement products to create a smoke-free home?

**QUESTIONS 7-10 FOR EVERYONE:**

7. Sometimes, whole communities declare they will keep their home smoke-free (for example, the ‘MyHouse’ initiative in Malaysia, and village based work in Indonesia).

Show the video clip below as an example – NOTE that this is about developing a completely smoke-free village. We are interested in supporting communities to create smoke-free homes only (stress that community members would not have to quit smoking completely!)

<https://www.dw.com/en/indonesias-non-smoking-neighborhood/video-63020609>

In this example from Indonesia, each house paints their door a bright colour when it becomes a smoke-free home. There are other approaches that can help to support communities to create a smoke-free home. For example, households may be given stickers and posters to put up on their door/in their house, and tips on how to create a smoke-free home [show the examples below, which can be laminated on card to pass around]

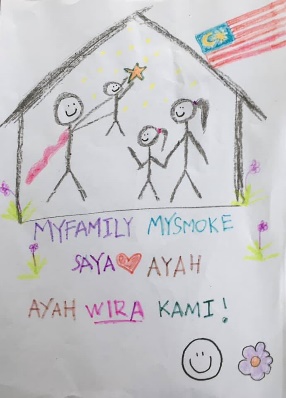

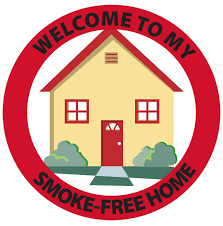

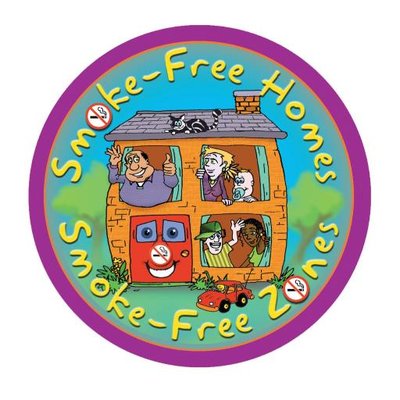

8. What do you think about the idea of supporting communities to create smoke-free homes? Could a community smoke-free home approach work here? If so, what might that look like? And who might be involved? (children, families, community leaders, schools, health clinics?)
9. Do you have any other thoughts on ways in which a) you, b) members of your family who smoke c) members of this community could be supported to create/maintain a smoke-free home? (for example, website resources, leaflets, support from health professionals, or any of the above suggestions discussed today in combination with each other)
10. Finally, if at any point a) you, b) someone in your family, c) a member of this community wanted to quit smoking, what support is available to support you/them with this? Are there local quit smoking services that you know of? (NEXT QUESTION FOR SMOKERS ONLY) Have you ever used them before/Would you consider using them? (Explore why/why not)

**Summary and close (5 mins)**

To summarise…. [briefly summarise key points of discussion…] Have I missed anything you feel is important? Does anyone have any other questions or comments before we finish?

We really appreciate your time today, and thank you for sharing your thoughts and opinions.
