## Supplementary file 2 for "Communities facilitating increasing smoke-free homes (CO-FRESH): co-developing an intervention with local stakeholders in Indonesia and Malaysia"

**Supplementary file 2: COFRESH WP3 INTERVIEW SCHEDULE**

*Interviews will be semi-structured; the order and phrasing of the questions will depend on the participant’s responses*.

**Introduction**

- I would like to know about your views on receiving the information from air quality feedback, any associated impacts (positive or negative) and to co-develop messaging for campaign development to promote implementation of Smoke-free home in the community.
- There are no right or wrong answers and your views and experiences are important.
- This session will take no longer than one hour.
- You can withdraw at any time from the interview, and you can choose not to answer certain questions.
- I would like to record the conversation for write-up purpose. The recording only accessible for research team members and your name and address will not be on the written-up transcript of your interview.
- I would like to use some of your words in my final report, presentations and articles for academic journals but it will be anonymised.
- Obtain informed consent from participant.

1. **Views on receiving personalised air quality feedback**
a. How did you feel when you first saw your personalised-feedback on air quality in your home? *(NOTE: responses may be different for men and women, so ask each individual in turn and then briefly summarise any differences/similarities in accounts)*
b. What do you think now about the information you have received on air quality in your home?
c. Did anything surprise you about the information you received? (*ask each individual in turn*) (PROMPT: Did you learn anything new? Were levels higher or lower than you expected them to be?)
d. Did anything worry you about the information you received? Any other negative impacts? (ask each individual in turn)
e. Are there any positive impacts of receiving this information? For you? Other members of the family? (PROMPT: Have you discussed or shared the air quality feedback with other members of the family/friends/visitors to the home? Have you kept any of the feedback materials (put them on the fridge for example)?
2. **Possible impacts on home smoking behaviours**
f. What difference, if any, do you think having this personalised feedback will make/made to you? Has it led to any changes in where you smoke? Why?
g. If you have made changes towards becoming a smoke-free home, where do you now smoke around the home?
h. How easy or difficult would it be for you to change when and where you smoke in smoke in the home? Why? (Note: Break these into separate ‘when’ and ‘where’ questions if easier, asking ‘why’ for both)
i. How easy/difficult would it be for you to create a smoke-free home? Why?
3. **Feedback on information provided**
j. How could the information you were given be improved? i.e. is it easy enough to understand, could it be presented differently etc? What about the graphs? Could they be displayed differently to make them easier to understand? Is there any additional information they would like to receive?
k. Could the information about the air quality measurement results be easily understood without an explanation from the research team?
4. **Wider Information on SHS harms**
l. Generally, do you feel like you have enough information about second-hand smoke harms to guide decisions about when and where you smoke? (probe on information regarding health harms and reasons for smoking outside in particular) If not, what information would you find helpful? Who should provide this information?
m. Knowing what you now know about air quality in your home, do you think others might benefit from personalised feedback on air quality? Why/Why not?
n. What is the single most important message that you would like others to know about second-hand smoke in the home? How would you communicate this message to others?
o. Is there anything else you would like to add about your experience of taking part in this work?
p. Any other general comments?
