## Supplementary file 3 for "Communities facilitating increasing smoke-free homes (CO-FRESH): co-developing an intervention with local stakeholders in Indonesia and Malaysia"

**Supplementary file 3: COFRESH training module content, materials and aims**

| **Sub-module** | **Title** | **Materials included** | **Aim** |
| --- | --- | --- | --- |
| 1 | CO-FRESH: What is the ‘Smoke Free Home Inititative’? | Handout and video | To inform understanding of what a smoke-free home means, why it is important, and the benefits of creating a smoke-free home. |
| 2 | Preparing to create a smoke-free home | Handout and video | To assist preparation for creating a smoke-free home. |
| 3 | Barriers and facilitators to creating a smoke-free home | Handout and video | To identify challenges associated with creating a smoke-free home and strategies to overcome these challenges. |
| 4 | Tips and resources to assist with creating a smoke-free home | Handout and video | To assist with facilitating creating a smoke-free home. |
