## Supplementary file 4 for "Communities facilitating increasing smoke-free homes (CO-FRESH): co-developing an intervention with local stakeholders in Indonesia and Malaysia"

**Supplementary file 3: WP1 interview schedule for pretesting of module content with health professionals - Indonesia**

Interviews will be semi-structured; the order and phrasing of the questions will depend on the participant’s responses.

**Introduction**

1. I would like to know about your views on the “smoke-free home” training module for health workers to promote implementation of Smoke-free home in the community.
2. There are no right or wrong answers and your views and experiences are important.
3. This session will take no longer than 1 hour.
4. You can withdraw at any time from the interview, and you can choose not to answer certain questions.
5. I would like to record the conversation for write-up purpose. The recording will only accessible to research team members and your name and address will not be on the written-up transcript of your interview.
6. We may like to use some of your words in my final report, presentations and articles for academic journals but this will be anonymised.
7. I would like to obtain informed consent from you if you agree to become a participant in this study.
8. I will provide you with the link to view the module online on the device that I have here.

**Topic Guide questions**

1. What is your overall impression of the smoke-free home training module for health care workers?
2. How relevant and important do you think the smoke-free home training module is for health care workers in promoting smoke-free environments?
3. What benefits did Health Workers get after reading this module? Did you gain any new knowledge after reading this module?
4. Could you share any specific strengths or weaknesses you have observed in the smoke-free home training module? In your opinion, what can we do to address these shortcomings?
5. What aspects of the smoke-free home training module do you think are most effective in promoting behavior change among patients?
6. Are there any areas where you believe the smoke-free home training module could be improved? How could we improve this area?
7. How confident do you feel in delivering the key messages and interventions outlined in the smoke-free home training module? Are there any areas/ part of the module you would find more difficult to deliver? Why?
8. What additional resources or support do you think would enhance the effectiveness of the smoke-free home training module?
