## Supplementary file 5 for "Communities facilitating increasing smoke-free homes (CO-FRESH): co-developing an intervention with local stakeholders in Indonesia and Malaysia"

**Supplementary file 4: WP1 interview schedule for pretesting of module content with health professionals - Malaysia**

INTERVIEW TOPIC GUIDELINES FOR HEALTHCARE WORKERS

QUALITATIVE INTERVIEW

COFRESH

This interview is semi-structured; the order and phrasing of the questions will depend on the participants' responses.

Introduction

1. Introduce Yourself (Interviewer)

2. Purpose: To gain insights from healthcare workers regarding the Smoke-Free Home Training Module aimed at promoting smoke-free homes in the community.

3. Clarification: There are no right or wrong answers to the questions provided; your views and experiences are important.

4. Duration: This session will not exceed 1 hour.

5. Voluntary Participation: You can withdraw at any time and may choose not to answer certain questions.

6. Recording: I would like to record the conversation for writing purposes. This recording will only be accessible to the research team, and your name and address will not be included in the interview transcript.

7. Use of Your Words: I would also like to use your responses in the final report, presentation, and academic journal articles, but they will be anonymized.

Before the Q&A Session

Ask the Healthcare Worker to Introduce Themselves:

a) Name, age, profession (doctor/nurse/Midwife Assistant)

b) educational background

c) Work experience

d) Do you treat patients in Felda around Kuala Kubu Bharu?

Questions

1. Have you thoroughly reviewed the Smoke-Free Home Training Module?

2. Do you provide advice about smoke-free homes to residents?

3. Do you offer information on how to quit smoking to residents?

4. What is your general impression of the Smoke-Free Home Training Module for healthcare workers?

5. In your opinion, how important and suitable is the Smoke-Free Home Training Module for healthcare workers?

6. What benefits do you perceive from reading this Smoke-Free Home Training Module for healthcare workers (e.g., quitting smoking, using air quality measurement tools)?

7. Can you specifically share any strengths and weaknesses you have noticed in the Smoke-Free Home Training Module?

8. Which aspects of the Smoke-Free Home Training Module do you think are effective in promoting behavior change among smokers?

9. Where in the Smoke-Free Home Training Module do you believe improvements can be made?

10. Do you feel confident in conveying the main messages and interventions outlined in the Smoke-Free Home Training Module?

11. What additional resources or support do you think are necessary to enhance the effectiveness of the Smoke-Free Home Training Module?

12. In your opinion, who do you think is qualified/suitable to be trained to explain the Smoke-Free Home Training Module?
