## Supplementary material for "Communities facilitating increasing smoke-free homes (CO-FRESH): co-developing an intervention with local stakeholders in Indonesia and Malaysia": Reflexivity statement

### **Supplementary file 6: Reflexivity Statement**

1. **How does this study address local research and policy priorities?**

As a result of high prevalences of adult male smoking, children in South-East Asia have a particularly high rates of exposure to second-hand smoke (SHS); 58% of children are exposed to second-SHS in Indonesia and 49% in Malaysia. Whilst reducing children’s exposure to SHS is a global public health priority, interventions to support families to create a smoke-free home have largely been developed in high income countries, focusing on smoking behaviour change at the individual or household level. These approaches have limited value in South-East Asian settings because of social and cultural norms regarding male smoking behaviour. A community-wide approach has been shown to hold promise, alongside training of allied health professionals. This study co-developed the CO-FRESH smoke-free homes intervention toolkit, which could be delivered in small village/town communities in Indonesia and Malaysia by healthcare professionals, community leaders and/or peers. Our focus on a community-based approach addresses local needs, preferences and research and policy priorities, working towards reducing exposure to second-hand smoke in both countries.

**2.	How were local researchers involved in study design?**

The first category of local researchers involved were those with extensive experience of involvement designing, conducting, leading, or organising international research on smoke-free homes (EZA and AZ in Malaysia, YSP, BSB and RSP in Indonesia). The second category of local researchers involved were those working as researchers, who were also directly involved in fieldwork (AWR in Indonesia, and AIA, NHM, IAMS, NSS, WTZ and SNAK in Malaysia). In addition, there were high-income country researchers with extensive experience of designing, conducting, leading, or organising international research collaborations involving low- and middle-income countries (SS, RO, IU).

3. **How has funding been used to support the local research team?**

This project was used to leverage funds to support 4 days per month time allocation for the Malaysian and Indonesian site co-I and leads, together with funds for a full-time research assistant and administrative assistant (Malaysia), and a full-time project manager and research assistant (Indonesia), for the life of the project. Small sums for travel within both countries, rental of community rooms for meetings and equipment were also included, and costs for hosting/travelling to a three day project meeting in Indonesia were included in the project budget.

4. **How are research staff who conducted data collection acknowledged?**

All research staff who conducted data collection are included as authors.

5. **Do all members of the research partnership have access to study data?**

All members of the partnership have access to data.

6. **How was data used to develop analytical skills within the partnership?**

Research Assistants in Malaysia who didn’t have previous experience of qualitative analysis attended capacity building training sessions which were led by RO and IU at the University of Stirling. The qualitative analysis process was undertaken with support from both University of Stirling team members. Research Assistants in Malaysia and Indonesia who didn’t have previous experience of measurement of indoor air quality attended a capacity building training session led by SS at the University of Stirling, who provided additional support regarding measurement and quantitative analysis throughout the study.

7. **How have research partners collaborated in interpreting study data?**

The three day in-person workshop held in Indonesia included sessions dedicated to discussion and interpretation of initial study data, alongside identification of areas for additional analysis. At the end of the workshop we agreed our key initial findings based on discussion of each data obtained in each workpackage.

8. **How were research partners supported to develop writing skills?**

All co-authors actively contributed to this paper, and Malaysian and Indonesian teams were also allocated specific sections of the paper to lead on, with early career researchers supported by the senior author(s) within their Institution.

9. **How will research products be shared to address local needs?**

This paper will be published as open access. We have developed a post-publication dissemination plan to distribute recommendations across a wide constituency, including in-country conference presentations, blogs and international webinars.

10. **How is the leadership, contribution and ownership of this work by LMIC researchers recognised within the authorship?**

Authors BSB worked as part of the senior authorship team in developing this manuscript, and we have specifically included his contribution as joint first author. The authorship team is predominantly based in Indonesia and Malaysia (12 LMIC authors and 3 HIC authors) and includes early, mid and senior LMIC researchers, and mid and senior HIC authors.

11. **How have early career researchers across the partnership been included within the authorship team?**

We have included all early career researchers involved in this research in Indonesia and Malaysia within the authorship team. They contributed to the fieldwork, analysis, write up of the study and in several cases attended the three day in-person workshop in Indonesia.

12. **How has gender balance been addressed within the authorship?**

Three authors are male (BSB, AIA, SS – one from each study team) and 12 authors are female (RO, AWR, RSP, NHM, IAMS, NSS, WTZ, SNAK, AZ, IU, EZA, YSP)

13. **How has the project contributed to training of LMIC researchers?**

Research funding leveraged as part of this project supported employment of junior researchers based in Indonesia and Malaysia. As outlined in response to question 6, Research Assistants in Malaysia who didn’t have previous experience of qualitative analysis attended capacity building training sessions which were led by RO and IU at the University of Stirling. Research Assistants in Malaysia and Indonesia who didn’t have previous experience of measurement of indoor air quality attended a capacity building training session led by SS at the University of Stirling.

14. **How has the project contributed to improvements in local infrastructure?**

This project has not directly contributed to improvements in local infrastructure.

15. **What safeguarding procedures were used to protect local study participants and researchers?**

The study was granted ethical approval by University of Stirling General University Ethical Panel (GUEP 2022 10527 7993, 25/10/2022), Universitas Gadjah Mada Faculty of Medicine, Public Health and Nursing Medical and Health Research Ethics Committee (KE/FK/0099/EC/2023) and the University Putra Malaysia Ethics Committee for Research involving Human Subjects (JKEUPM-2023-047). Safeguarding was an integral part of our overarching research ethics.
